# Twenty-four hour activity-count behavior patterns associated with depressive symptoms: A big data-machine learning approach

**DOI:** 10.1101/2023.08.09.23293905

**Authors:** Saida Salima Nawrin, Hitoshi Inada, Haruki Momma, Ryoichi Nagatomi

**Author notes:** Department of Biochemistry & Cellular Biology, National Center of Neurology and Psychiatry, Kodaira, Tokyo, Japan (Hitoshi Inada). Corresponding author (HI) (RN). These authors contributed equally to this work (Saida Salima Nawrin, Hitoshi Inada).

## Abstract

Depression is a global burden with profound personal and economic consequences. Previous studies have reported that the amount of physical activity is associated with depression. However, the relationship between the temporal patterns of physical activity and depressive symptoms is poorly understood. We hypothesize that the temporal patterns of daily physical activity could better explain the association of physical activity with depressive symptoms. To address the hypothesis, we investigated the association between depressive symptoms and daily dominant activity behaviors based on 24-hour temporal patterns of physical activity. We found that evening dominant behavior was positively associated with depressive symptoms compared to morning dominant behavior as the control group. Our results might contribute to monitoring and identifying individuals with latent depressive symptoms, emphasizing the importance of nuanced activity patterns and their probability of assessing depressive symptoms effectively.

## Introduction

Depression is a mood disorder that causes persistent sadness and loss of interest in activities one used to enjoy [1]–[4]. Depression is a prevalent mental health condition affecting about 280 million people worldwide and responsible for more than 47 million disability-adjusted life years in 2019 [5][6]. It is also a leading cause of disability globally and is associated with premature mortality from other illnesses [7] and suicide [8]. Depression also imposes substantial economic burdens. In 2010, it was estimated that approximately $US210.5 billion was spent annually in the US alone on lost work productivity and medical treatment associated with depression [9]–[11]. Moreover, the costs experienced an increase of 12.9% between 2010 and 2018 [11]. The manifestation of depressive symptoms is an important sign of the onset of depression [2][4]. Therefore, understanding the prevalence of depressive symptoms within the population is vital for effective public health planning [12]. It enables officials and policymakers to gauge the magnitude of the issue and allocate necessary resources for prevention, diagnosis, and treatment. Furthermore, highlighting the prevalence of depressive symptoms can reduce the stigma surrounding mental health problems by demonstrating that it is a widespread and manageable condition [13].

Physical activity is a factor associated with depression [14]. A recent observational study of US adults found that every additional hour of light-intensity physical activity decreased the chances of being depressed by 20%; even moderate-to-vigorous physical activity was linked to a lower risk of depression [15]. Another study mentioned that each additional hour of light activity per day between ages 12 and 16 reduced depression scores by 8-11% at age 18 [16]. Similarly, higher physical activity levels, especially among women, are associated with a reduced likelihood of having depression [17]. Consistent with these observations, studies involving interventions have demonstrated that all modes of physical activity effectively reduce depressive symptoms, although high-intensity physical activity shows more significant improvements in reducing mild to moderate symptoms of depression, anxiety, and psychological distress in diverse adult populations.[18].

While recent investigations challenge the notion that a higher volume or intensity of physical activity is beneficial for depressive symptoms [19][20], some studies have indicated a potential association between the timing of physical activity and depressive symptoms [21][22], possibly due to the shift in the circadian rhythm experienced by individuals with depression [23]. The shift in the circadian rhythm could result in changes in physical, mental, and behavioral patterns throughout the 24-hour cycle, leading to a change in preferred times for engaging in activities among depressive populations [24][25]. Therefore, the physical activity pattern in terms of timing could be a helpful and vital parameter in understanding the association between physical activity and depressive symptoms.

Several studies have explored the relationship between physical activity patterns, specifically in timing, and the association with depressive symptoms [26]. For example, higher depressive symptoms are associated with late-night activity around 1:30 a.m. [27]. There was a tendency towards a later timing of activity peak in the acute depression group [21]. Similarly, participants with depression exhibited notably lower physical activity from 7 a.m. to 10 p.m. (daytime) compared to the healthy control group [28]. By contrast, a previous study has reported that individuals with depression and anxiety exhibited lower overall activity levels but did not show a specific association between the timing of activity and the presence or severity of depressive symptoms [29]. No differences in the timing of physical activity between depressed and non-depressed individuals were found, even when participants receiving antidepressant treatment were excluded from the analysis [30]. Considering these reports, the association between physical activity patterns and depressive symptoms remains inconsistent.

The inconsistency of the association between depressive symptoms and physical activity patterns, including timing or phase, might be due to the difficulty of establishing a standardized procedure to categorize physical activity patterns. Previous studies can be attributed to the limitations in capturing the behavior of physical activity based on the temporal shape of the activity for each day [21][27]–[30]. By aggregating data from multiple days, the unique temporal shape of each day’s activity is diluted, potentially masking the associations with depressive symptoms [31]. These temporal shapes could be critical because depressive individuals have more fragmented physical activity throughout the day because of distorted circadian rhythm [31][32]. Thus, the temporal shape of physical activity could be a valuable marker for detecting depressive symptoms. [32][33]

Recently, machine learning (ML) has gained increasing prominence in the field of epidemiology due to its potential for effective disease prediction and patient care [33]**–**[36]. The ML is also applied to detecting physical activities in daily behaviors such as sleeping, sitting, walking, ascending stairs, descending stairs, and running from big data obtained from wearable devices or smartphones [37]–[49]. These studies focused on the specific type or the mode of the activity rather than the repetitive patterns or the behavior of the physical activity. Since raw physical activity data is often complex and multi-dimensional, involving various parameters like duration, intensity, type, and context, analyses by ML, which enables us to handle complexity and relationships within the data, are beneficial, even when non-linear relationships or interactions between variables exist [50]–[52]. Traditional statistical methods may struggle with such complexity and may require simplifications or assumptions that can lead to less accurate categorization [50]–[52]. In addition, another advantage of the ML approach is to enable us to measure accurate physical activity over a long period, possibly leading to figuring out physical activity patterns, which could be challenging to handle and unrecognized by manual assessments [33]**–**[36]

In this study, we investigated the relationship between activity behaviors, defined as the probability of temporal physical activity patterns, and depressive symptoms using big data of the accelerometer from the National Health and Nutrition Examination Survey (NHANES) 2011-2012 [53] by time-series analysis with ML [54]. Here, the physical activity-count behaviors were determined by the probability of the physical activity-counting patterns, which are classified as temporal shapes based on physical activity-counting within 24 hours. The physical activity-count behaviors would provide us with insights into the specific type of physical activity pattern that each individual engages through a week, while the physical activity-counting patterns reflect how an individual’s activity is distributed throughout the day, based on rhythms or lifestyle. We classified four distinct activity-count behaviors based on the dominant activity pattern. Individuals with the Evening dominant behavior had a higher prevalence of depressive symptoms than those with the Morning dominant behavior (as a reference), even though they showed similar total activity count. These findings could highlight the importance of distribution in activity behaviors for understanding the relationship between physical activity and depressive symptoms. Recognizing the patterns and variability of physical activity might contribute to finding a behavioral context for manifesting depressive symptoms.

## Results

### Demographic characteristics of activity-count behavior

Twenty-four-hour activity-counting patterns and daily activity-count behaviors were determined using the ML procedure reported previously [54]. Five 24-hour activity-counting patterns all day (AD), morning (M), evening (E), bi-phasic (BP), and irregular morning (IM) and 4 clusters of daily activity-count behaviors (AD dominant, M dominant, AD+M dominant, and E dominant) were identified (Figure 1, Figure 2 and Supplementary Tables S1-S3). Participants with depressive symptom data (n = 4242) were extracted from the clustered data (n = 6613) and used in subsequent analyses. Table 1 presents the demographic characteristics of each activity-count behavior. Out of 4242 participants, M dominant behavior held the maximum number of participants (n = 1444, 34.04%), followed by AD+M dominant (n = 1075, 25.34%), AD dominant (n = 1012, 23.86%), and E dominant (n = 711, 16.76%) behaviors. For age, M dominant is the oldest adult (55.82 ± 16.43 years), followed by AD+M dominant (47.47 ± 17.60 years), AD dominant (44.68 ± 18.52 years), and E dominant (34.75 ± 15.76 years) (p < 0.0001). BMI does not show a significant difference among the activity-count behavior (p = 0.1284), and the BMI is highest for the AD+M dominant (29.18 ± 7.03) and lowest for the AD dominant (28.49 ± 7.09). AD dominant has the lowest percentage of male participants (44.47%), but E dominant showed the highest number of male participants (55.27%), M dominant (52.22%), and AD+M dominant (49.12%) followed. The work situation also differs between activity-count behaviors (p < 0.0001). The total activity is highest in AD dominant (13114.39±3606.71), followed by M dominant (12844.19±4140.83), AD+M dominant (11960.01±4292.37), and E dominant (11544.57±4926.13). E dominant showed the highest prevalence of depressive symptoms (15.75%), followed by AD+M dominant (8.56%), AD dominant (8.40%), and M dominant (6.99%) (p < 0.0001).

**Figure 1.**
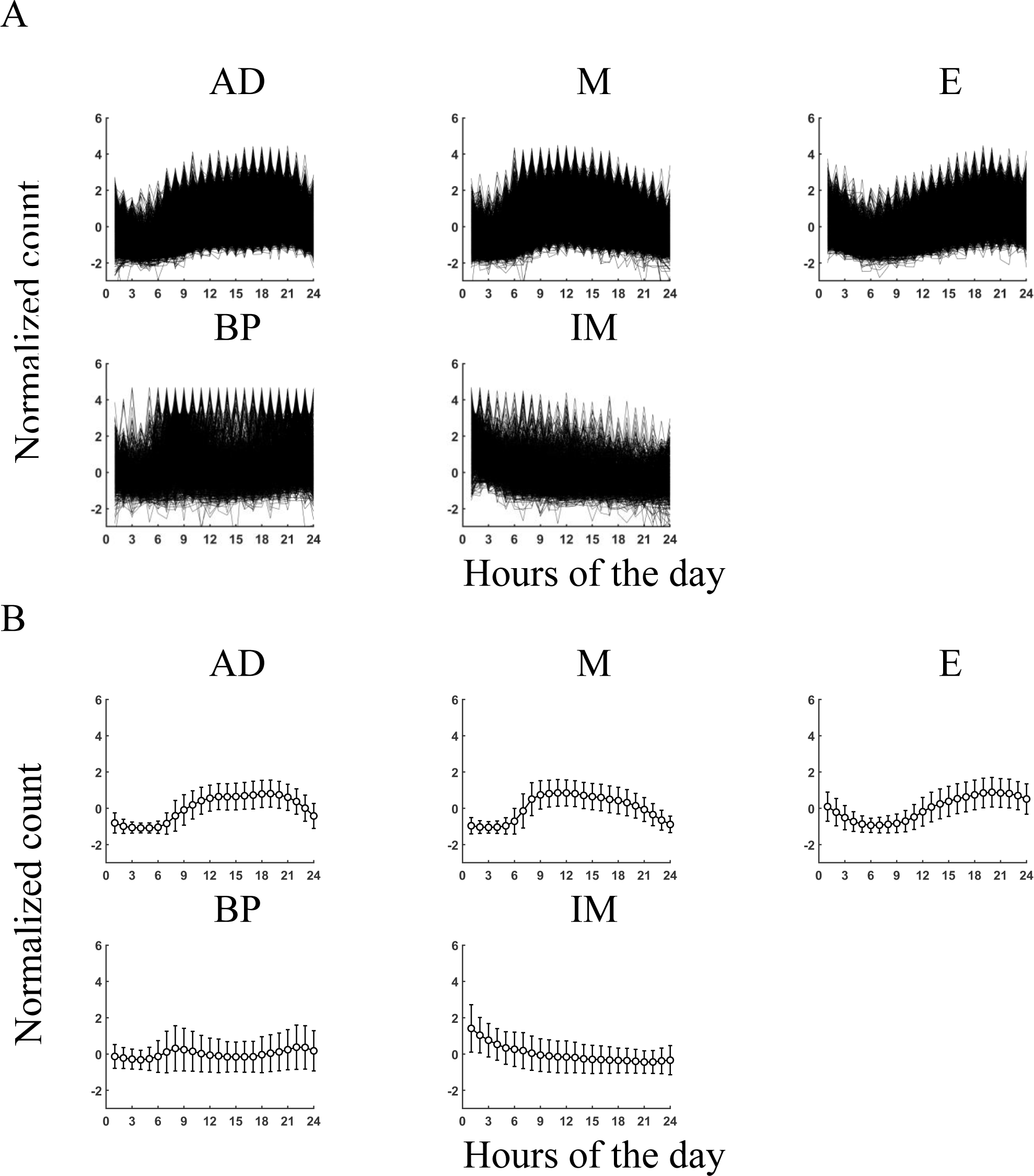
Five 24-hour physical activity patterns. A. Individual plots of standardized activity count for each pattern. B. Plots of mean values with standard deviation (SD) for each pattern. AD, All-day; M, Morning; E, Evening; BP, Bi-phasic; IM, Irregular morning.

**Figure 2.**
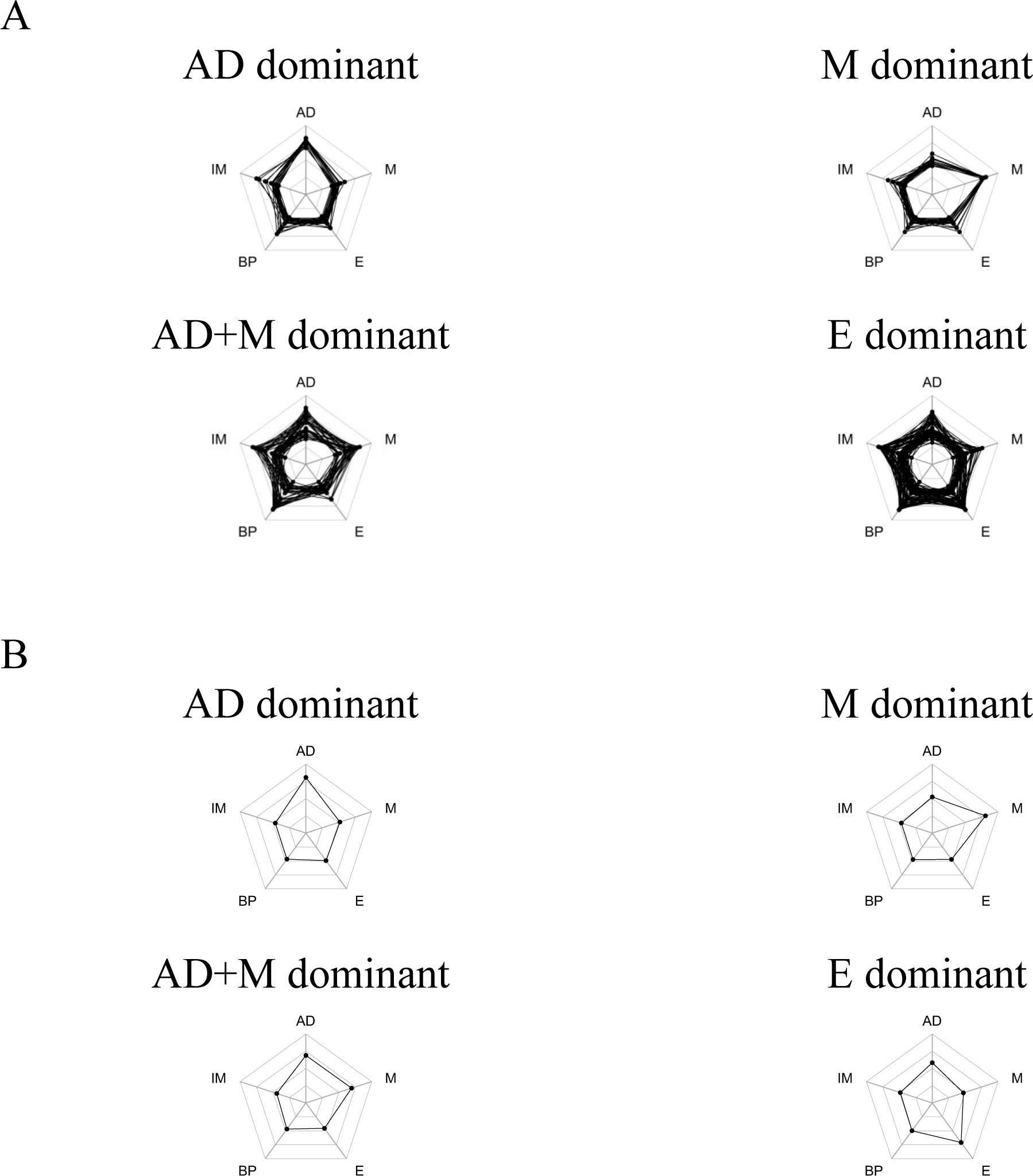
Four daily behavioral patterns. A. Individual spider plots of the probability of 24-hour physical activity patterns. B. Spider plots of mean values for each pattern. AD, All-day; M, Morning; AD+M, All day+Morning; E, Evening.

**Table 1.**
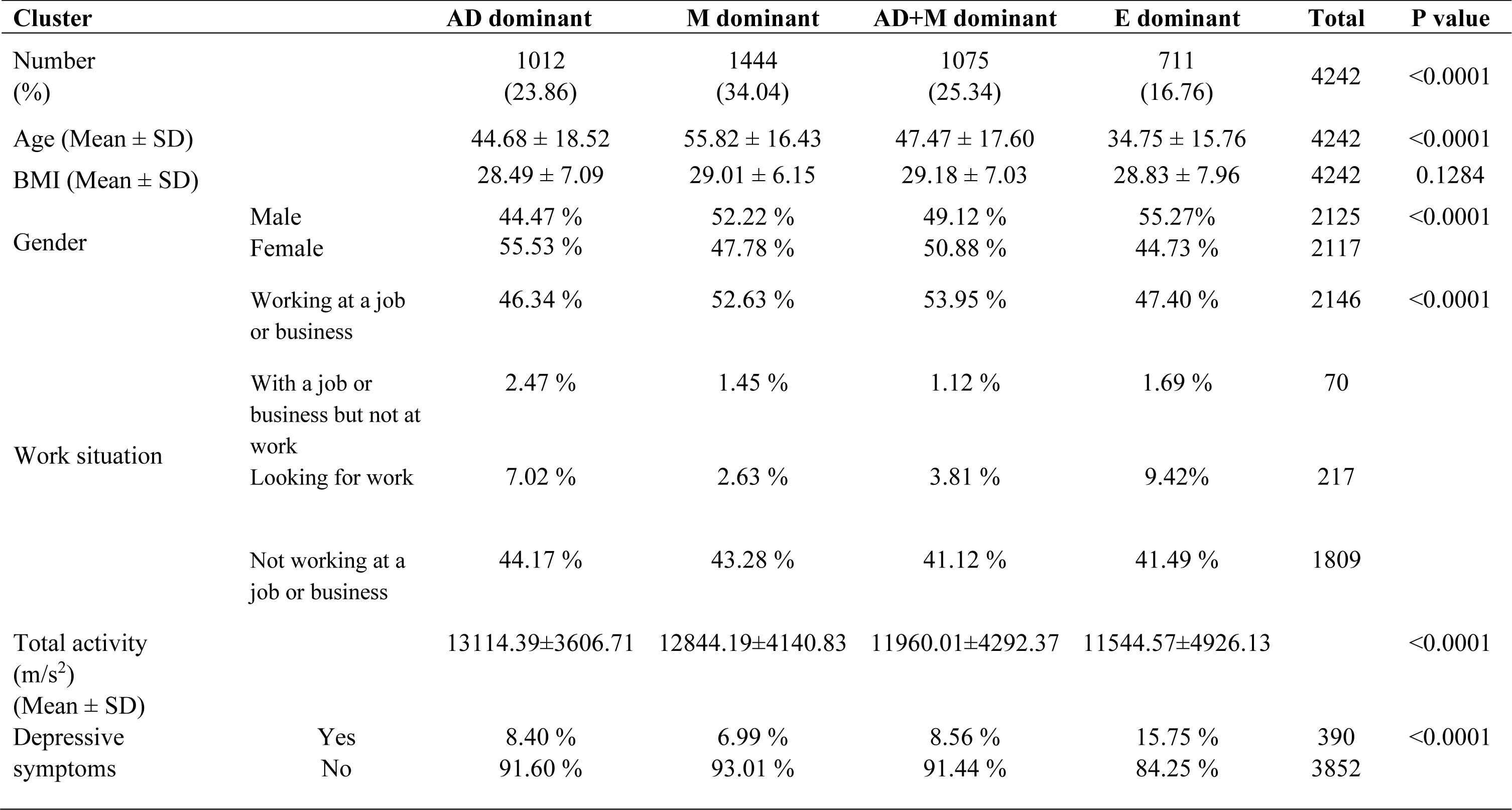
Demographic characteristics of the clusters.

### Relationship between activity-count behavior and depressive symptoms

Table 2 represents the odds ratios (ORs) and 95% confidence intervals (CIs) for the depressive symptoms based on activity-count behaviors. The reference group is the M dominant behavior, with the lowest percentage of individuals with depressive symptoms. In the crude model (Model 1), the OR for depressive symptoms was 2.48 (95% CI: 1.86– 3.30) for the E dominant behavior, 1.24 (95% CI: 0.92–1.67) for the AD+M dominant behavior, and 1.21 (95% CI: 0.90–1.64) for the AD dominant behavior. A significant association between E dominant behavior and depressive symptoms was observed after adjusting for covariates, including age, BMI, gender, work situation, and total activity (Model 2 to 6). No strong correlation was observed among these covariates (Supplementary Figure S1). A significant difference was observed after adjusting for each confounding factor (age, BMI, gender, work situation, and total activity). E dominant behavior remains associated with a higher prevalence of depressive symptoms compared to M dominant behavior (p < 0.0001), as shown in Supplementary Table S4. These results suggest that the association between E dominant behavior and depressive symptoms is robust against these confounding factors.

**Table 2.** Odd ratios for depressive symptoms by activity-count behavior.

|  | Behavior |  |  |  |  |
| --- | --- | --- | --- | --- | --- |
|  | M dominant | AD dominant | AD+M dominant | E dominant | p-value |
| Model 1 | 1.00 (ref) | 1.21(0.90,1.64) | 1.24(0.92,1.67) | 2.48(1.86,3.30) | <0.0001 |
| Model 2 | 1.00 (ref) | 1.32(0.96,1.79) | 1.32(0.98,1.78) | 2.88(2.11,3.96) | <0.0001 |
| Model 3 | 1.00 (ref) | 1.31(0.96,1.78) | 1.28(0.95,1.73) | 2.80(2.04,3.83) | <0.0001 |
| Model 4 | 1.00 (ref) | 1.24(0.91,1.69) | 1.25(0.93,1.69) | 2.82(2.05,3.87) | <0.0001 |
| Model 5 | 1.00 (ref) | 1.09(0.79,1.50) | 1.19(0.88,1.62) | 2.30(1.66,3.20) | <0.0001 |
| Model 6 | 1.00 (ref) | 1.09(0.79,1.50) | 1.17(0.86,1.59) | 2.21(1.58,3.09) | <0.0001 |

## Discussion

In this study, we first characterized physical activity-counting patterns by examining the temporal shape of physical activity that reflects how an individual’s activity is distributed throughout 24 hours. Then, we defined physical activity-count behaviors by identifying the dominant physical activity-counting patterns, showing the repetitive feature of the physical activity that individuals engage in throughout the week. This procedure was achieved using unsupervised ML techniques. The current study identified five physical activity-counting patterns and four physical activity-count behaviors. Based on the identified physical activity-count behaviors, we revealed that individuals exhibiting E dominant (Evening dominant) behavior had a higher prevalence of depressive symptoms compared to those with M dominant (Morning dominant) behavior, irrespective of total activity count. This association was confirmed after adjusting for potential confounding factors. We also found that individuals with a higher prevalence of depressive symptoms have higher day-to-day variability.

Recently, studies using ML to analyze a huge data set of physical movements appeared with potential applications in epidemiology, public health, and human behavioral science [33]**–**[36]. The ML is applied to identify or categorize behaviors such as sleeping, lying, sitting, standing, walking, ascending or stairs, running, and other routine activities like biking, household chores, and yoga [37]–[49]. However, few studies have directly extracted or categorized temporal physical activity patterns by ML but could not differentiate between the temporal physical activity pattern and the intensity [55] or the total amount of physical activity [56]. Our study focused on categorizing each 24-hour activity-count into several clusters directly and identifying the behavioral tendencies of each individual based on the probability of dominant activity patterns. Our approach based on the data-driven analysis could be more straightforward, insightful, and beneficial to handle temporal patterns and dominant behaviors of human physical activity directly, although a relatively large complete data set is required.

Previous studies showed various results regarding the association between depressive symptoms and physical activity patterns, such as the timing of activity [27][28], consistent with our result that a specific temporal behavioral pattern (E-dominant) is associated with depressive symptoms. In these reports, the higher depression severity was linked to increased activity late at night in the calculated 24-hour activity pattern [27] and the depressed participants had significantly lower physical activity counts during the daytime compared to healthy controls [28]. In contrast, the other observational and case-control studies have indicated no association between depressive symptoms and activity timing [29][30]. Different classifications of the activity patterns due to the distinct procedures might result in inconsistent conclusions since these observational and case-control studies did not observe or detect the evening or nighttime activity patterns, possibly leading to no association of physical activity patterns with depressive symptoms.

In our results, the E pattern showed the highest activity between 6 p.m. and 9 p.m., and the E dominant behavior showed a higher prevalence of depressive symptoms. The association between E dominant behavior and the depressive symptom might reflect a disruption of circadian rhythm, which could play a crucial role in regulating mood and sleep-wake cycles since disruptions in the circadian rhythms, such as delayed sleep-wake timing, have been associated with increased risk of depressive symptoms and mood disorders [57]. Alternatively, individuals with depression may reduce daytime activity and compensate for total physical activity by increasing their evening activity when they feel energetic [58][59]. The latter possibility might explain our result in which E dominant (risk group) and M dominant (control group) behavior showed a similar total count activity.

Many previous studies reported that there is a connection between depressive symptoms and low physical activity levels [21][28][29]. These findings suggested that total physical activity would be significant for the association with depressive symptoms. For instance, individuals with depression had lower 24-hour physical activity levels compared to controls without depression, and those with acute depression exhibited a marginal significance towards later timing of daily activity peaks, particularly in the evening [21]. Similarly, individuals with depression displayed significantly lower physical activity levels between 7 a.m. and 10 p.m. compared to healthy controls, considering both timing and activity levels [28]. Additionally, the presence and severity of depressive and anxiety disorders were associated with reduced overall daily activity levels, but no significant association was found with the timing of activity [29]. In our study, E-dominant behavior exhibited a total activity count similar to AD+M-dominant behavior (Table 1). However, only E-dominant behavior was found to be associated with depressive symptoms compared to M-dominant behavior (table 2). On the other hand, the AD dominant behavior has a higher amount of activity count compared to the M dominant behavior (Table 1), but the AD dominant behavior is not associated with depressive symptoms (Table 2). These results suggest that the physical activity behavior based on the dominant physical activity pattern might be a more significant parameter than the total amount of physical activity to explain the association with depressive symptoms in a particular procedure.

We also examined the day-to-day variability of activity patterns within different activity behaviors (Supplementary Table S3) and their relationship with depressive symptoms. We found that AD dominant, M dominant, and AD+M dominant behaviors exhibit the least day-to-day variability, with nearly 80% of their dominant activity patterns corresponding to their respective behaviors. However, the E dominant behavior shows considerably more variability, as only 49% of E patterns align with E dominant behavior, while the rest are distributed among other patterns (Supplementary Table S3). In this analysis, we observed that individuals with higher day-to-day variability (E dominant behavior) had a higher prevalence of depressive symptoms compared to those with less day-to-day variability in their activity behavior (AD dominant, M dominant, and AD+M dominant behavior) (Table 1). These findings indicate that the variability and temporal shape of activity patterns may play a significant role in the relationship between physical activity and depressive symptoms. This underscores the importance of considering these factors when seeking to understand the connection between physical activity and depressive symptoms.

Recent research has yielded valuable insights into the multifaceted influences on depressive symptoms across different life stages and factors, such as age, BMI, gender, and employment. Age was associated with a gradual increase in depressive symptoms, especially among older individuals [60]. Furthermore, a study on the relationship between depressive symptoms and BMI anticipated that overweight or obese individuals would experience more severe depressive symptoms, regardless of their racial background [61]. There are gender differences in depressive disorder, where women are more likely to have a higher prevalence of depression than men [62]. Unemployment could be associated with onset of clinical major depression [63]. Our study uncovered a higher prevalence of depressive symptoms among individuals engaged in E-dominant behavior compared to those with M-dominant behavior, even after adjusting for confounding factors above. Our finding provides insight into the contribution of the physical activity pattern to understanding depressive syndrome, emphasizing the importance of individualized assessment of physical activity behavior to understand depressive symptoms.

A major limitation of the current study is that the causal relationship between activity-count behavior and depressive symptoms could not be established because of the cross-sectional design. In addition, the assessment of depressive symptoms was used by self reported questionnaires. Although PHQ-9 used in this study has high sensitivity and specificity [64]**–**[66], we did not precisely diagnose the depression. However, our study would provide a possible impact of physical activity patterns rather than the total amount of physical activity on the relationship with depressive symptoms. Evaluating the physical activity/behavioral patterns, such as temporal changes or probability, may lead to valuable insights into the association between physical activity and depressive symptoms. Our findings also have a potential benefit to developing software for wearable devices alerting or reminding a risk to prevent the progression of depressive symptoms. These findings might have implications for improving the management and prevention of depressive symptoms in clinical and community settings.

## Methods

### Activity-counting data

National Health and Nutrition Examination Survey (NHANES) 2011-2012 data was used for the analysis [53]. Participants were asked to wear a wrist-worn ActiGraph model GT3X+ accelerometer for seven consecutive days, 24 hours a day, starting on the day of their examination. The device was water-resistant, triaxial, and could detect the magnitude of acceleration at 80 Hz sampling intervals. The device was worn on the non-dominant wrist, and participants were instructed to wear it continuously except when they needed to temporarily remove it.

### Depression data

The NHANES 2011-2012 uses the PHQ-9 questionnaire to measure depression. The Patient Health Questionnaire (PHQ-9) is a commonly used screening tool to assess the presence and severity of depression. It consists of nine questions about the frequency of symptoms of depression experienced over the past 2 weeks. Each question has four response categories: “not at all,” “several days,” “more than half the days,” and “nearly every day.” Each response category is assigned a point ranging from 0 to 3, with 0 indicating that the symptom was not present and 3 indicating that the symptom was present nearly every day. The scores for each item are then summed to give a total score ranging from 0 to 27. The final follow-up question in the PHQ-9 assesses the overall impairment caused by depressive symptoms. This question asks the patient to rate the degree to which their symptoms have caused problems in their daily life, including work, school, or relationships. Overall, the PHQ-9 provides a simple and reliable method for identifying individuals experiencing depression and assessing the severity of their symptoms. It is widely used in clinical practice and research to screen for depression and monitor the response to treatment. PHQ-9 score > or =10 had a sensitivity of 88% and a specificity of 88% for major depression [64]**–**[66].

### Software and data processing

MATLAB (2021a, MathWorks, MA) and Python (3.11.0) were used for data processing and visualization.

### Twenty-four hour activity-counting pattern and daily activity-count behaviors

In this analysis, the minute-level physical activity data from the NHANES 2011-2012 dataset is utilized. Only participants with 1440 minutes of data for 7 complete days (totaling 6613 participants) are included to ensure data completeness. The data is then converted into a standardized format (mean = 0 and SD = 1) for each 24 hours, resulting in a dataset comprising 46291 days of 24-hour standardized data. To cluster the 24-hour physical activity patterns, unsupervised ML using the tslearn Python package is employed [67]. The probability of each 24-hour physical activity pattern was obtained for each participant to figure out dominant daily behavioral patterns. Vectors of the probability of each 24-hour physical activity pattern (with five values) were used to cluster the daily behavioral patterns by unsupervised ML using a K-means clustering algorithm with MATLAB. Four clusters of daily behavioral patterns were obtained. Five clusters of 24-hour physical activity patterns and four behavioral patterns are shown in Figure 1 and Figure 2.

### Depression measurement

The study used the PHQ-9 questionnaire to measure depression. Out of the total 6613 participants, 4,242 participants provided complete answers to all nine questions, and they have the complete dataset for Age, Gender, BMI, and Work situation. The questionnaire provides a total score ranging from 0 to 27, with a threshold score of 10 being used to determine depression [68]. Participants who scored less than 10 were labeled as not having depression, while those who scored 10 or more were labeled as having depression. Out of 4242 participants, 390 participants have depression, and 3852 participants have no depression.

### Statistical analysis

The statistical analyses were performed using JMP software (version 16.2.0). For the demographic analysis, we reported descriptive statistics such as means and standard deviations (SD) for continuous variables and percentages for categorical variables stratified by activity-count behavior. To compare differences in continuous variables, we used ANOVA, while chi-square tests were employed for categorical variables across different activity-count behaviors.

To investigate the relationship between activity-count behavior and depressive symptoms, we conducted a multivariable logistic regression analysis. Depressive symptoms were the dependent variable, and activity-count behavior was the independent variable. We calculated odds ratios (ORs) and 95% confidence intervals (CIs). Initially, we examined the crude model (Model 1), which assessed the association between activity-count behavior and depressive symptoms without considering any confounding factors. Subsequently, we introduced one confounding factor at a time, progressing from Model 2 to Model 6.

In order to assess the impact of residual confounding, we conducted a sensitivity analysis. This analysis aimed to determine the influence of age, BMI, gender, work situation, and total activity on the association between activity-count behavior and depressive symptoms. We performed logistic regression analyses similar to the previous models, adding only one individual confounding factor in each model. We used the Wald test to assess the significance of the association in both cases. We also checked the correlations between each individual variable. We considered statistical significance as p < 0.05 for all analyses.

## Data Availability

All data produced are available online at

https://wwwn.cdc.gov/nchs/nhanes/continuousnhanes/default.aspx?BeginYear=2011

## Supporting information

**S1 Fig. Correlation coefficients.**

Work situation and age showed a very weak positive correlation. Work situation and total activity showed a very weak negative correlation. Other variables showed no correlation among them.

**S1 Table. Number of days for each activity counting pattern**

**S2 Table. Number of participants for behavior.**

**S3 Table. Proportion of activity counting patterns in each behavior.**

**S4 Table. Sensitivity analysis (Odd ratios for depressive symptoms by activity behavior)**

## Acknowledgments

We would like to thank all the participants who have contributed to the experiment. A pioneering research support grant from Tohoku University partially supported this project. The funding body has no role in the study design, data analysis, or manuscript submission.

## Author contributions

SSN and HI analyzed the data. SSN, HI, and HM wrote the manuscript. HI and RN designed and supervised the research. All authors revised the manuscript and approved the final manuscript.

## Data availability

https://wwwn.cdc.gov/nchs/nhanes/continuousnhanes/default.aspx?BeginYear=2011

## Competing interest

The authors declare no competing interests.

## Notes

### Competing Interest Statement

The authors have declared no competing interest.

### Funding Statement

This study was partially funded by
JST SPRING
grant number: JPMJSP2114

### Author Declarations

The study used (or will use) ONLY openly available human data that were originally located at: https://wwwn.cdc.gov/nchs/nhanes/continuousnhanes/default.aspx?BeginYear=2011

### Summary of Updates

The revised version exclusively employs unsupervised machine learning instead of utilizing a mixed approach that combines supervised and unsupervised techniques. The rationale for employing a machine learning approach is more explicitly articulated in this version.

## References

[1] S. P. Chand and H. Arif, Depression. StatPearls Publishing, 2023. Accessed: Oct. 09, 2023. [Online]. Available: https://www.ncbi.nlm.nih.gov/books/NBK430847/

[2] “Psychiatry.org - What Is Depression?” Accessed: Oct. 09, 2023. [Online]. Available: https://www.psychiatry.org/patients-families/depression/what-is-depression

[3] “What is Depression? | SAMHSA.” Accessed: Oct. 09, 2023. [Online]. Available: https://www.samhsa.gov/mental-health/depression

[4] “Depressive disorder (depression).” Accessed: Oct. 09, 2023. [Online]. Available: https://www.who.int/news-room/fact-sheets/detail/depression

[5] C. Abbafati et al., “Global burden of 369 diseases and injuries in 204 countries and territories, 1990–2019: a systematic analysis for the Global Burden of Disease Study 2019,” The Lancet, vol. 396, no. 10258, pp. 1204–1222, 2020, doi: 10.1016/S0140-6736(20)30925-9.

[6] “Depressive disorder (depression).” [Online]. Available: https://www.who.int/news-room/fact-sheets/detail/depression

[7] E. R. Walker, R. E. McGee, and B. G. Druss, “Mortality in mental disorders and global disease burden implications a systematic review and meta-analysis,” JAMA Psychiatry, vol. 72, no. 4, pp. 334–341, 2015, doi: 10.1001/jamapsychiatry.2014.2502.

[8] V. Patel et al., “Addressing the burden of mental, neurological, and substance use disorders: Key messages from Disease Control Priorities, 3rd edition,” The Lancet, vol. 387, no. 10028, pp. 1672–1685, 2016, doi: 10.1016/S0140-6736(15)00390-6.

[9] W. E. Broadhead, D. G. Blazer, L. K. George, and C. K. Tse, “Depression, Disability Days, and Days Lost From Work in a Prospective Epidemiologic Survey,” JAMA: The Journal of the American Medical Association, vol. 264, no. 19, pp. 2524–2528, 1990, doi: 10.1001/jama.1990.03450190056028.

[10] C. J. L. M. and A. D. Lopez and It, “Evidence-Based Health Policy-Lessons from the Global Burden of Disease Study,” Science (1979), vol. 274, pp. 1–4, 1996.

[11] P. E. Greenberg et al., “The Economic Burden of Adults with Major Depressive Disorder in the United States (2010 and 2018),” Pharmacoeconomics, vol. 39, no. 6, pp. 653–665, 2021, doi: 10.1007/s40273-021-01019-4.

[12] CDC, “CDC Promotes Public Health Approach to Address Depression Among Older Adults.” Accessed: Oct. 09, 2023. [Online]. Available: https://www.cdc.gov/aging/pdf/cib_mental_health.pdf

[13] P. W. Corrigan, S. B. Morris, P. J. Michaels, J. D. Rafacz, and N. Rüsch, “Challenging the public stigma of mental illness: a meta-analysis of outcome studies,” Psychiatr Serv, vol. 63, no. 10, pp. 963–973, Oct. 2012, doi: 10.1176/APPI.PS.201100529.

[14] R. K. DIshman, C. P. McDowell, and M. P. Herring, “Customary physical activity and odds of depression: A systematic review and meta-analysis of 111 prospective cohort studies,” British Journal of Sports Medicine, vol. 55, no. 16. BMJ Publishing Group Ltd and British Association of Sport and Exercise Medicine, pp. 926–934, Aug. 01, 2021. doi: 10.1136/bjsports-2020-103140.

[15] P. D. Loprinzi, “Objectively measured light and moderate-to-vigorous physical activity is associated with lower depression levels among older US adults,” Aging Ment Health, vol. 17, no. 7, pp. 801–805, Sep. 2013, doi: 10.1080/13607863.2013.801066.

[16] A. Kandola, G. Lewis, D. P. J. Osborn, B. Stubbs, and J. F. Hayes, “Depressive symptoms and objectively measured physical activity and sedentary behaviour throughout adolescence: a prospective cohort study,” Lancet Psychiatry, vol. 7, no. 3, pp. 262–271, Mar. 2020, doi: 10.1016/S2215-0366(20)30034-1.

[17] P. D. Loprinzi and B. J. Cardinal, “Interrelationships among physical activity, depression, homocysteine, and metabolic syndrome with special considerations by sex,” Prev Med (Baltim), vol. 54, no. 6, pp. 388–392, Jun. 2012, doi: 10.1016/J.YPMED.2012.03.016.

[18] B. Singh et al., “Effectiveness of physical activity interventions for improving depression, anxiety and distress: an overview of systematic reviews,” Br J Sports Med, p. bjsports-2022-106195, 2023, doi: 10.1136/bjsports-2022-106195.

[19] H. Shimamoto, M. Suwa, and K. Mizuno, “Relationships between depression, daily physical activity, physical fitness, and daytime sleepiness among japanese university students,” Int J Environ Res Public Health, vol. 18, no. 15, 2021, doi: 10.3390/ijerph18158036.

[20] F. Guo, Y. Tian, F. Zhong, C. Wu, Y. Cui, and C. Huang, “Intensity of physical activity and depressive symptoms in college students: Fitness improvement tactics in youth (fityou) project,” Psychol Res Behav Manag, vol. 13, pp. 797–811, 2020, doi: 10.2147/PRBM.S266511.

[21] O. Minaeva et al., “Level and timing of physical activity during normal daily life in depressed and non-depressed individuals,” Transl Psychiatry, vol. 10, no. 1, 2020, doi: 10.1038/s41398-020-00952-w.

[22] A. Wirz-Justice, “Biological rhythm disturbances in mood disorders,” Int Clin Psychopharmacol, vol. 21, no. SUPPL. 1, Feb. 2006, doi: 10.1097/01.yic.0000195660.37267.cf.

[23] M. A. Quera Salva, S. Hartley, F. Barbot, J. C. Alvarez, F. Lofaso, and C. Guilleminault, “Circadian rhythms, melatonin and depression,” Curr Pharm Des, vol. 17, no. 15, pp. 1459–1470, Jun. 2011, doi: 10.2174/138161211796197188.

[24] A. Germain and D. J. Kupfer, “Circadian rhythm disturbances in depression,” Human Psychopharmacology: Clinical and Experimental, vol. 23, no. 7, pp. 571–585, Oct. 2008, doi: 10.1002/HUP.964.

[25] W. Bechtel, “Circadian rhythms and mood disorders: Are the phenomena and mechanisms causally related?,” Front Psychiatry, vol. 6, no. AUG, p. 153068, Aug. 2015, doi: 10.3389/FPSYT.2015.00118/BIBTEX.

[26] V. Gianfredi et al., “Association between Daily Pattern of Physical Activity and Depression: A Systematic Review,” Int J Environ Res Public Health, vol. 19, no. 11, p. 6505, Jun. 2022, doi: 10.3390/IJERPH19116505/S1.

[27] N. Banihashemi et al., “Quantifying the effect of body mass index, age, and depression severity on 24-h activity patterns in persons with a lifetime history of affective disorders,” BMC Psychiatry, vol. 16, no. 1, Sep. 2016, doi: 10.1186/S12888-016-1023-2.

[28] E. A. Wolff, F. W. Putnam, and R. M. Post, “Motor activity and affective illness. The relationship of amplitude and temporal distribution to changes in affective state,” Arch Gen Psychiatry, vol. 42, no. 3, pp. 288–294, 1985, doi: 10.1001/ARCHPSYC.1985.01790260086010.

[29] S. Difrancesco et al., “Sociodemographic, health and lifestyle, sampling, and mental health determinants of 24-hour motor activity patterns: Observational study,” J Med Internet Res, vol. 23, no. 2, pp. 1–14, 2021, doi: 10.2196/20700.

[30] N. Lorenz, J. Spada, C. Sander, S. G. Riedel-Heller, and U. Hegerl, “Circadian skin temperature rhythms, circadian activity rhythms and sleep in individuals with self-reported depressive symptoms,” J Psychiatr Res, vol. 117, pp. 38–44, Oct. 2019, doi: 10.1016/j.jpsychires.2019.06.022.

[31] R. Brady, W. J. Brown, M. Hillsdon, and G. I. Mielke, “Patterns of Accelerometer-Measured Physical Activity and Health Outcomes in Adults: A Systematic Review,” Med Sci Sports Exerc, vol. 54, no. 7, pp. 1155–1166, 2022, doi: 10.1249/MSS.0000000000002900.

[32] M. De Feijter, D. Kocevska, M. A. Ikram, and A. I. Luik, “The bidirectional association of 24-h activity rhythms and sleep with depressive symptoms in middle-aged and elderly persons,” 2023.

[33] T. L. Wiemken and R. R. Kelley, “Machine Learning in Epidemiology and Health Outcomes Research,” Annu Rev Public Health, vol. 41, pp. 21–36, Apr. 2020, doi: 10.1146/ANNUREV-PUBLHEALTH-040119-094437.

[34] S. Rose, “Intersections of machine learning and epidemiological methods for health services research,” Int J Epidemiol, vol. 49, no. 6, pp. 1763–1770, Jan. 2021, doi: 10.1093/IJE/DYAA035.

[35] M. Javaid, A. Haleem, R. Pratap Singh, R. Suman, and S. Rab, “Significance of machine learning in healthcare: Features, pillars and applications,” International Journal of Intelligent Networks, vol. 3, pp. 58–73, Jan. 2022, doi: 10.1016/J.IJIN.2022.05.002.

[36] A. J. Hamilton et al., “Machine learning and artificial intelligence: applications in healthcare epidemiology,” Antimicrobial Stewardship & Healthcare Epidemiology: ASHE, vol. 1, no. 1, pp. 1–6, Oct. 2021, doi: 10.1017/ASH.2021.192.

[37] C. Dobbins and R. Rawassizadeh, “Towards Clustering of Mobile and Smartwatch Accelerometer Data for Physical Activity Recognition,” Informatics 2018, Vol. 5, Page 29, vol. 5, no. 2, p. 29, Jun. 2018, doi: 10.3390/INFORMATICS5020029.

[38] H. He, Y. Tan, and W. Zhang, “A wavelet tensor fuzzy clustering scheme for multi-sensor human activity recognition,” Eng Appl Artif Intell, vol. 70, pp. 109–122, Apr. 2018, doi: 10.1016/J.ENGAPPAI.2018.01.004.

[39] P. Jones et al., “Towards a Portable Model to Discriminate Activity Clusters from Accelerometer Data,” Sensors 2019, Vol. 19, Page 4504, vol. 19, no. 20, p. 4504, Oct. 2019, doi: 10.3390/S19204504.

[40] P. J. Jones et al., “FilterK: A new outlier detection method for k-means clustering of physical activity,” J Biomed Inform, vol. 104, p. 103397, Apr. 2020, doi: 10.1016/J.JBI.2020.103397.

[41] M. Kheirkhahan, A. Chakraborty, A. A. Wanigatunga, D. B. Corbett, T. M. Manini, and S. Ranka, “Wrist accelerometer shape feature derivation methods for assessing activities of daily living,” BMC Med Inform Decis Mak, vol. 18, no. 4, pp. 1–13, Dec. 2018, doi: 10.1186/S12911-018-0671-1/FIGURES/9.

[42] P. Lago and S. Inoue, “Comparing feature learning methods for human activity recognition: Performance study in new user scenario,” 2019 Joint 8th International Conference on Informatics, Electronics and Vision, ICIEV 2019 and 3rd International Conference on Imaging, Vision and Pattern Recognition, icIVPR 2019 with International Conference on Activity and Behavior Computing, ABC 2019, pp. 118–123, May 2019, doi: 10.1109/ICIEV.2019.8858548.

[43] I. Machado, R. Gomes, H. Gamboa, and V. Paixão, “Human Activity Recognition from Triaxial Accelerometer Data Feature Extraction and Selection Methods for Clustering of Physical Activities”, doi: 10.5220/0004749801550162.

[44] I. P. Machado, A. Luísa Gomes, H. Gamboa, V. Paixão, and R. M. Costa, “Human activity data discovery from triaxial accelerometer sensor: Non-supervised learning sensitivity to feature extraction parametrization,” Inf Process Manag, vol. 51, no. 2, pp. 204–214, Mar. 2015, doi: 10.1016/J.IPM.2014.07.008.

[45] A. Nguyen, D. Moore, and I. McCowan, “Unsupervised clustering of free-living human activities using ambulatory accelerometry,” Annual International Conference of the IEEE Engineering in Medicine and Biology - Proceedings, pp. 4895–4898, 2007, doi: 10.1109/IEMBS.2007.4353437.

[46] D. Van Kuppevelt, J. Heywood, M. Hamer, S. Sabia, E. Fitzsimons, and V. Van Hees, “Segmenting accelerometer data from daily life with unsupervised machine learning,” PLoS One, vol. 14, no. 1, p. e0208692, Jan. 2019, doi: 10.1371/JOURNAL.PONE.0208692.

[47] D. Wang, L. Liu, X. Wang, and Y. Lu, “A novel feature extraction method on activity recognition using smartphone,” Lecture Notes in Computer Science (including subseries Lecture Notes in Artificial Intelligence and Lecture Notes in Bioinformatics), vol. 9998 LNCS, pp. 67–76, 2016, doi: 10.1007/978-3-319-47121-1_6/TABLES/1.

[48] X. Wang, Y. Lu, D. Wang, L. Liu, and H. Zhou, “Using jaccard distance measure for unsupervised activity recognition with smartphone accelerometers,” Lecture Notes in Computer Science (including subseries Lecture Notes in Artificial Intelligence and Lecture Notes in Bioinformatics), vol. 10612 LNCS, pp. 74–83, 2017, doi: 10.1007/978-3-319-69781-9_8/FIGURES/2.

[49] C. Y. Yong, R. Sudirman, N. H. Mahmood, and K. M. Chew, “Motion Classification Using Proposed Principle Component Analysis Hybrid K-Means Clustering *,” no. 5, pp. 25–30, 2013, doi: 10.4236/eng.2013.55B006.

[50] D. Bzdok, N. Altman, and M. Krzywinski, “Points of Significance: Statistics versus machine learning,” Nat Methods, vol. 15, no. 4, pp. 233–234, Apr. 2018, doi: 10.1038/NMETH.4642.

[51] H. S. R. Rajula, G. Verlato, M. Manchia, N. Antonucci, and V. Fanos, “Comparison of Conventional Statistical Methods with Machine Learning in Medicine: Diagnosis, Drug Development, and Treatment,” Medicina (B Aires), vol. 56, no. 9, pp. 1–10, Sep. 2020, doi: 10.3390/MEDICINA56090455.

[52] C. Ley, R. K. Martin, A. Pareek, A. Groll, R. Seil, and T. Tischer, “Machine learning and conventional statistics: making sense of the differences,” Knee Surgery, Sports Traumatology, Arthroscopy, vol. 30, no. 3, pp. 753–757, Mar. 2022, doi: 10.1007/S00167-022-06896-6/FIGURES/1.

[53] “Centers for Disease Control and Prevention (CDC). National Center for Health Statistics (NCHS). National Health and Nutrition Examination Survey Data. Hyattsville, MD: U.S. Department of Health and Human Services, Centers for Disease Control and Prevention, [2011-2012].” Accessed: Jul. 15, 2023. [Online]. Available: https://wwwn.cdc.gov/nchs/nhanes/continuousnhanes/default.aspx?BeginYear=2011

[54] S. S. Nawrin, H. Inada, H. Momma, and R. Nagatomi, “Clustering temporal step-counting patterns for 24 hours with machine learning revealed potential heterogeneity in the categorization by a traditional tertile procedure,” bioRxiv, 2023, doi: 10.1101/2023.06.04.543652.

[55] M. Niemelä et al., “Intensity and temporal patterns of physical activity and cardiovascular disease risk in midlife,” 2019, doi: 10.1016/j.ypmed.2019.04.023.

[56] M. Aqeel et al., “Temporal Physical Activity Patterns are Associated with Obesity in U.S. Adults,” Prev Med (Baltim), vol. 148, p. 106538, Jul. 2021, doi: 10.1016/J.YPMED.2021.106538.

[57] A. Wirz-Justice and R. H. Van Den Hoofdakker, “Sleep deprivation in depression: What do we know, where do we go?,” Biol Psychiatry, vol. 46, no. 4, pp. 445–453, 1999, doi: 10.1016/S0006-3223(99)00125-0.

[58] L. M. Trotti, “Waking up is the hardest thing I do all day: Sleep inertia and sleep drunkenness,” Sleep Med Rev, vol. 35, p. 76, Oct. 2017, doi: 10.1016/J.SMRV.2016.08.005.

[59] S. F. Smagula, C. S. Capps, and R. T. Krafty, “Evaluating the timing of differences in activity related to depression symptoms across adulthood in the United States,” J Affect Disord, vol. 284, p. 64, Apr. 2021, doi: 10.1016/J.JAD.2021.01.069.

[60] A. Fiske, M. Gatz, and N. L. Pedersen, “Depressive Symptoms and Aging: The Effects of Illness and Non-Health-Related Events,” The Journals of Gerontology: Series B, vol. 58, no. 6, pp. P320–P328, Nov. 2003, doi: 10.1093/GERONB/58.6.P320.

[61] N. Badillo, M. Khatib, P. Kahar, and D. Khanna, “Correlation Between Body Mass Index and Depression/Depression-Like Symptoms Among Different Genders and Races,” Cureus, vol. 14, no. 2, Feb. 2022, doi: 10.7759/CUREUS.21841.

[62] M. Piccinelli and G. Wilkinson, “Gender differences in depression: Critical review,” The British Journal of Psychiatry, vol. 177, no. 6, pp. 486–492, 2000, doi: 10.1192/BJP.177.6.486.

[63] B. J. Jefferis et al., “Associations between unemployment and major depressive disorder: evidence from an international, prospective study (the predict cohort),” Soc Sci Med, vol. 73, no. 11, pp. 1627–1634, Dec. 2011, doi: 10.1016/J.SOCSCIMED.2011.09.029.

[64] R. L. Spitzer, K. Kroenke, and J. B. W. Williams, “Validation and utility of a self-report version of PRIME-MD: The PHQ Primary Care Study,” J Am Med Assoc, vol. 282, no. 18, pp. 1737–1744, Nov. 1999, doi: 10.1001/jama.282.18.1737.

[65] K. Kroenke and R. L. Spitzer, “The PHQ-9: A new depression diagnostic and severity measure,” Psychiatr Ann, vol. 32, no. 9, pp. 509–515, 2002, doi: 10.3928/0048-5713-20020901-06.

[66] K. Kroenke, R. L. Spitzer, and J. B. W. Williams, “The PHQ-9: Validity of a brief depression severity measure,” J Gen Intern Med, vol. 16, no. 9, pp. 606–613, 2001, doi: 10.1046/j.1525-1497.2001.016009606.x.

[67] R. Tavenard et al., “Tslearn, A Machine Learning Toolkit for Time Series Data,” Journal of Machine Learning Research, vol. 21, pp. 1–6, 2020.

[68] S. F. Smagula et al., “Association of 24-Hour Activity Pattern Phenotypes with Depression Symptoms and Cognitive Performance in Aging,” JAMA Psychiatry, vol. 79, no. 10, pp. 1023–1031, 2022, doi: 10.1001/jamapsychiatry.2022.2573.

