## Supplementary figures and images for "Twenty-four hour activity-count behavior patterns associated with depressive symptoms: A big data-machine learning approach"

### Supplemental figure S1

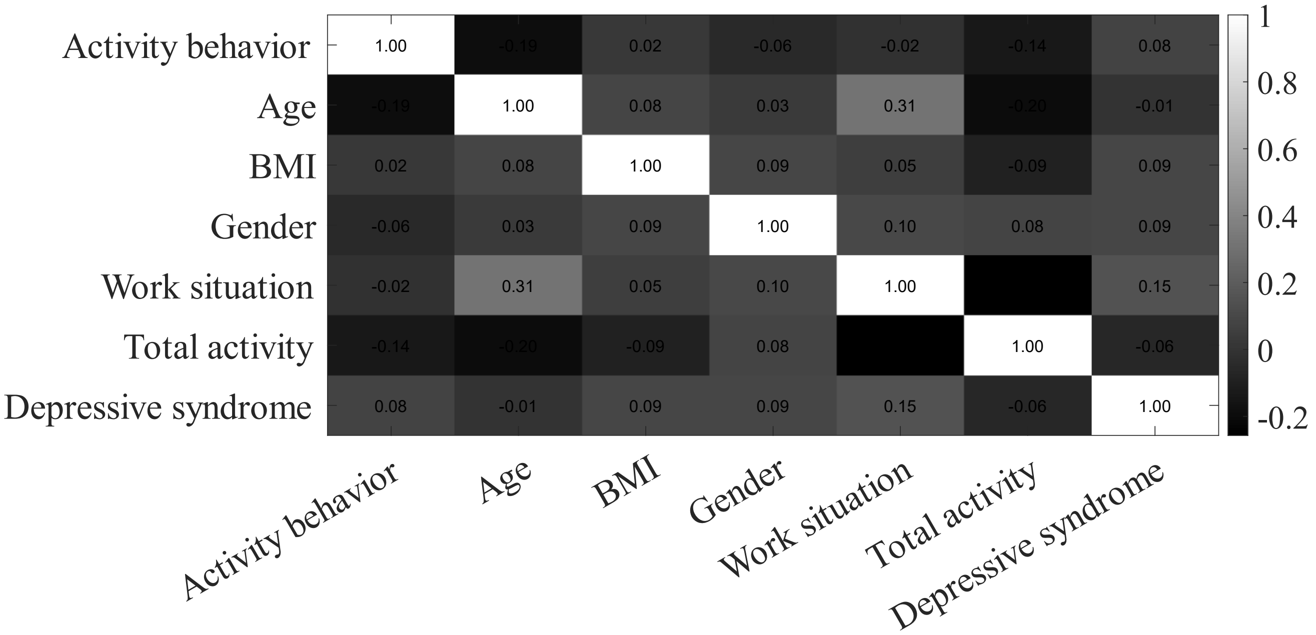
