## Supplemental table S1-S4 for "Twenty-four hour activity-count behavior patterns associated with depressive symptoms: A big data-machine learning approach"

**Table S1. Number of days for each activity-counting pattern**

| Activity patterns | No of days | % of days | Daily activity count (mean ± SD) |
|---|---|---|---|
| AD | 18980 | 41.00 | 14786.06±5454.00 |
| M | 19156 | 41.38 | 14004.09±5405.45 |
| E | 5033 | 10.87 | 12586.48±5551.76 |
| BP | 1932 | 4.17 | 4640.34±5755.53 |
| IM | 1190 | 2.57 | 7016.28±7419.17 |
| Total | 46291 | 100 | |

**Table S2. Number of participants for behavior**

| Activity-count Behavior | No of participants | % of participants | Total activity count (mean ± SD) |
|---|---|---|---|
| AD dominant | 1792 | 27.09 | 14996.28±4636.84 |
| M dominant | 1964 | 29.69 | 13628.09±4594.38 |
| AD+M dominant | 1855 | 28.05 | 13307.74±5063.63 |
| E dominant | 1002 | 15.15 | 11589.80±5254.22 |
| Total | 6613 | 100 | |

**Table S3. Proportion of activity-counting patterns in each behavior**

| | Activity-counting pattern probability (Mean ± SD) | | | | |
|---|---|---|---|---|---|
| Activity-count Behavior | AD | M | E | BP | IM |
| AD dominant | 0.7900±0.1281 | 0.1201±0.0192 | 0.0668±0.1016 | 0.0140±0.0499 | 0.0091±0.0395 |
| M dominant | 0.1269±0.1114 | 0.8341±0.1303 | 0.0135±0.0505 | 0.0175±0.0580 | 0.0081±0.0371 |
| AD+M dominant | 0.4342±0.1340 | 0.4396±0.1168 | 0.0433±0.0782 | 0.0594±0.1264 | 0.0236±0.0707 |
| E dominant | 0.2407±0.1772 | 0.0677±0.0958 | 0.4916±0.2396 | 0.1061±0.1793 | 0.0940±0.1604 |

**Table S4: Sensitivity analysis (Odd ratios for depressive symptoms by activity behavior)**

| Model | Behavior pattern | | | | P value |
|---|---|---|---|---|---|
| | M dominant | AD dominant | AD+M dominant | E dominant | p-value |
| Model 1 | 1.00 (ref) | 1.21(0.90,1.64) | 1.24(0.92,1.67) | 2.48(1.86,3.30) | <0.0001 |
| Model 2 | 1.00 (ref) | 1.32(0.96,1.79) | 1.32(0.98,1.78) | 2.88(2.11,3.96) | <0.0001 |
| Model 3 | 1.00 (ref) | 1.23(0.91,1.66) | 1.22(0.91,1.64) | 2.48(1.87,3.31) | <0.0001 |
| Model 4 | 1.00 (ref) | 1.16(0.86,1.57) | 1.22(0.90,1.64) | 2.56(1.92,3.41) | <0.0001 |
| Model 5 | 1.00 (ref) | 1.18(0.87,1.60) | 1.27(0.95,1.72) | 2.52(1.89,3.38) | <0.0001 |
| Model 6 | 1.00 (ref) | 1.23(0.92,1.67) | 1.19(0.89,1.61) | 2.35(1.76,3.14) | <0.0001 |

Notes
Model 1: Activity behavior (Crude)
Model 2: Activity behavior, Age
Model 3: Activity behavior, BMI
Model 4: Activity behavior, Gender
Model 5: Activity behavior, work situation
Model 6: Activity behavior, total activity
